# The development of competency frameworks in healthcare professions: a scoping review

**DOI:** 10.1101/19003475

**Authors:** Alan M. Batt, Walter Tavares, Brett Williams

## Abstract

**Background:** Competency frameworks serve various roles including outlining characteristics of a competent workforce, facilitating mobility, and analysing or assessing expertise. Given these roles and their relevance in the health professions, we sought to understand the methods and strategies used in the development of existing competency frameworks.

**Methods:** We applied the Arksey and O’Malley framework to undertake this scoping review. We searched six electronic databases (MEDLINE, CINAHL, PsycINFO, EMBASE, Scopus, and ERIC) and three grey literature sources (greylit.org, Trove and Google Scholar) using keywords related to competency frameworks. We screened studies for inclusion by title and abstract, and we included studies of any type that described the development of a competency framework in a healthcare profession. Two reviewers independently extracted data including study characteristics. Data synthesis was both quantitative and qualitative.

**Results:** Among 5,710 citations, we selected 190 for analysis. The majority of studies were conducted in medicine and nursing professions. Literature reviews and group techniques were conducted in 116 studies each (61%), and 85 (45%) outlined some form of stakeholder deliberation. We observed a significant degree of diversity in methodological strategies, inconsistent adherence to existing guidance on the selection of methods, who was involved, and based on the variation we observed in timeframes, combination, function, application and reporting of methods and strategies, there is no apparent gold standard or standardised approach to competency framework development.

**Conclusions:** We observed significant variation within the conduct and reporting of the competency framework development process. While some variation can be expected given the differences across and within professions, our results suggest there is some difficulty in determining whether methods were fit-for-purpose, and therefore in making determinations regarding the appropriateness of the development process. This uncertainty may unwillingly create and legitimise uncertain or artificial outcomes. There is a need for improved guidance in the process for developing and reporting competency frameworks.

## Introduction

As individual health professions evolve, identification of competencies describing required knowledge, skills, attitudes and other characteristics (KSAOs) for effective professional practice are needed by professionals, educators, and regulators (Campion et al. 2011; Gonczi et al. 1990; Palermo et al. 2017). Identifying these competencies ensures that healthcare professions are well defined, promotes competent workforces, facilitates assessment, facilitates professional mobility, and helps to analyze and evaluate the expertise of the profession and the professional (Baer 1986; Campion et al. 2011; Eraut 1994; Gonczi et al. 1990; Heywood et al. 1992; ten Cate 2005; Whiddett and Hollyforde 1999; Winter and Maisch 2005; World Health Organization 2005). The CanMEDS framework, the ACGME Outcomes project, and the entry-level registered nurse practice competencies are examples of frameworks that have been used in these ways (Black et al. 2008; J. Frank et al. 2015; Swing 2007). Given the stakes that such frameworks hold for educators, learners, regulators, health professions and healthcare broadly, development guidelines have been created (See Table 1).

**Table 1.** Summary of existing framework development guidance

| Author/Year | Summary of guidance |
| --- | --- |
| Heywood 1992 | 1. Examine the existing information (government reports, studies undertaken by the profession, curriculum documents etc.)<br>2. Choose a combination of techniques (professions will need to choose a combination of techniques that address a range of practical and theoretical issues appropriate to the profession concerned)<br>a. The chosen methods should analyse the functions/activities/roles of the profession and the attributes of individuals<br>b. Methods should be practical and cost-effective<br>c. Methods should be acceptable to the profession<br>d. The more important the purpose of the framework, the more that validity needs to be assured using a combination of methods<br>3. Apply the methods<br>a. professions need to have developed a nationally representative steering group prior to commencing a project<br>4. Continuing consultation (widespread discussion of the project within the profession as a whole, both while it is being undertaken and after the development) |
| Roe 2002 | 1. Conduct occupational or job analysis<br>2. Perform competence analysis (including KSAOs)<br>3. Undertake competence modelling<br>4. Test the competence model |
| Marelli 2005 | 1. Define the Objectives<br>a. Why is there a need to develop a competency model?<br>b. What is the unit of analysis?<br>c. What is the relevant timeframe?<br>d. How will the competency model be applied?<br>e. Obtain the Support of a Sponsor<br>2. Develop and Implement a Communication and Education Plan<br>3. Plan the Methodology<br>a. Select the sample – use multiple groups, focus on high performers, identify desirable characteristics, select a representative sample<br>b. Select data collection methods – Use at least two methods that are |
|  | complementary. Suggests literature review as a preliminary approach. Suggests consideration of focus groups, interviews, surveys, observation, work logs, and competency menus.<br>4. Plan the Data Recording and Analysis<br>a. Identify the Competencies and Create the Competency Model<br>b. Define the job<br>c. Identify the competencies<br>5. Assemble the competency model<br>a. Review by subject matter experts<br>b. Develop behavioural examples<br>6. Apply the Competency Model<br>7. Evaluate and Update the Competency Model |
| Kwan 2016 | 1. Select the EPA topic<br>2. Develop the EPA content by collecting data from participants using focus group and individual interviews<br>3. Draft the EPAs based on analysis of collected data<br>4. Seek feedback on the draft EPAs from the participants and other stakeholders<br>5. Refine and finalise the EPAs based on feedback |
| Moerkamp and Onstenk 1991 in Klink and Boon 2002 | 1. Identify developments in the profession<br>2. Identify tasks of professionals<br>3. Identify competencies<br>4. Draft curriculum<br>5. Design or revise curriculum |
| Whiddett and Hollyforde, 1999; Whiddett and Hollyforde, 2003 | • Framework must be relevant to all those who may benefit from its use<br>• It must meet the needs of a wide range of possible applications<br>• Involve the people who will be affected by the framework in its development<br>• Keep people informed about what is happening and why during the development process<br>• Create competencies that are relevant<br>• Maintain a quality standard<br>Sequence of general stages:<br>• Get buy in from key people<br>• Clarify the purpose<br>• Plan the project<br>• Put together data gathering and analysis team<br>• Choose analysis techniques<br>• Gather data – the type of data collected will be influenced by the intended purpose of the competency framework<br>• Prepare for analysis<br>• Analyze data<br>• Draft competency framework<br>• Validate the competencies<br>• Revise and finalize the competencies<br>• Launch the framework |
| Lucia and Lepsinger 1999 | 1. Lay the groundwork<br>a. Determine objectives and scope<br>b. Clarify implementation goals and standards<br>c. Develop an action plan<br>d. Identify individuals at various performance levels<br>2. Develop the model<br>a. Determine data collection methodology<br>b. Collect data |
|  | c. Direct observation of incumbents<br>d. Develop an interim competency model<br>3. Finalize and validate<br>a. Test the competency model<br>b. Analyze the new data and refine the model<br>c. Validate the competency model<br>d. Finalize the model |
| Ten Cate 2005 | Competencies should be specific, comprehensive, durable, trainable, measurable, related to professional activities and connected to other competencies |
| Child and Shaw 2019 | Developers should consider three arguments when developing a competency framework in order to align the process with the intended use:<br>1. Binary vs continuum – is the framework going to be used to make a competent versus not-competent argument? Or is it to be used in a developmental manner (learning) manner?<br>2. Atomistic vs holistic – atomistic, checklist type competency frameworks can result in reductionist approaches to complex tasks. Holistic frameworks can face challenges when attempts are made to operationalize them due to lack of detail.<br>3. Context-specific vs Context-general – the intended generalisability and adaptability of a framework beyond particular contextual boundaries should help to determine the degree of contextual specificity incorporated into the development process. |
| Mansfield 2000 | 1. What HR application should we include in the initial model building project?<br>2. What will the key users of the model need from it?<br>3. How should key stakeholders be involved?<br>4. How extensive should the data collection be?<br>5. How should we balance research with intuitive approaches?<br>6. What format of behavioural descriptors will best suit the application?<br>7. How do we plan to accommodate additional, future competency models? |
| Campion et al 2011 | 1. Consider organizational context competency<br>2. Link competency models to organizational goals and objectives<br>3. Start at the top<br>4. Use rigorous job analysis methods to develop competencies<br>5. Consider future-oriented job requirements<br>6. Use additional unique methods<br>7. Define the anatomy of a competency<br>8. Define levels of proficiency on competencies<br>9. Use organizational language<br>10. Include both fundamental (cross-job) and technical (job-specific) competencies<br>11. Use competency libraries<br>12. Achieve the proper level of granularity (number of competencies and amount of detail)<br>13. Use diagrams, pictures, and heuristics to communicate competency models to employees<br>14. Use organizational development techniques to ensure competency modeling acceptance and use<br>15. Maintain the currency of competencies over time |

The development of competency frameworks requires strategies to capture and represent the complexity associated with healthcare practice. This complexity can emerge in a number of ways. For example, regional or contextual variability, unique practice patterns, the role and attributes of individuals and individuals within teams, and other interacting competencies make practice in multiple contexts possible (Bordage and Harris 2011; Garavan and McGuire 2001; Heywood et al. 1992; Hodges and Lingard 2012; Knapp and Knapp 1995; Lingard 2012; Makulova et al. 2015; Roe 2002). The nature of clinical practice can also be difficult to define or understand fully (Garavan and McGuire 2001; Mendoza 1994). The role of tacit knowledge in professional practice for example, can be difficult to represent - that is, there can be a disconnect between the personal knowledge of professionals which becomes embedded in their practice, and the publicly accessible knowledge base of the profession (Collin 1989; Eraut 1994). Competence and its component parts are often inconsistently understood or defined and attributed multiple meanings depending on context (Hay-McBer 1996; Spencer and Spencer 1993; ten Cate and Scheele 2007). Other difficulties may include shifts in patient demographics or societal expectations, the role of technology, and changes in organizational structures (Duong et al. 2017; Jacox 1997; Whiddett and Hollyforde 1999). As such, any attempt to represent professional practice must contend with these challenges, which leaves developers with decisions on how best to make those choices (Garavan and McGuire 2001; Shilton et al. 2001).

Given this inherent complexity in capturing and accurately representing the features of a health profession, a variety of approaches may be employed. Influencing issues such as practicality, efficiency, and what might be deemed acceptable to the profession may have a role (see Table 1). While there is no guidance on what specific methods to use, when to use them, or how to use them, there is consensus that in order to increase the validity and utility of competency frameworks a combination of approaches may be necessary, akin to a process of triangulation (Heywood et al. 1992; Klink and Boon 2002; Kwan et al. 2016; Marrelli et al. 2005). However, feasibility, the complexity of practice, and access to appropriate stakeholders may prompt developers to prioritise aspects of the developmental and validation process (Marrelli et al. 2005; Whiddett and Hollyforde 1999).

These challenges may result in variable or uncertain outcomes that may be of limited validity and utility (Lester 2014; Shilton et al. 2001). This in turn may inaccurately represent the profession, or represent it in unintended ways, and inappropriately impact downstream dependent systems such as policy/standards development, accreditation and curriculum. Given the activity related to the development of competency frameworks in many health professions, little attention has been paid to the development process. Despite existing guidelines, the complexity associated with different professional practices may lead some to enact development activities differently. This emphasis on actual developmental processes, in the context of existing but perhaps incomplete or inadequate guidelines, is the focus of our study. Understanding these activities may provide insights into how these processes shape eventual outcomes and their validity and/or utility, and provide insights into what may hold value for the refinement of existing guidelines. As such, the primary objective of our study is to understand the way in which health professions develop competency frameworks and then to consider these activities against existing guidance.

## Methods

### Design

We conducted a scoping review, which enabled us to identify, map and present an overview of a heterogeneous body of literature (Arksey and O’Malley 2005; Munn et al. 2018). We deemed a scoping review to be appropriate given our interest in identifying key characteristics of competency framework development as well as potential knowledge or practice gaps (Munn et al. 2018). We employed Arksey and O’Malley’s (2005) five-stage framework which included (1) identifying the research question, (2) identifying relevant studies, (3) refining the study selection criteria, (4) collecting relevant data from each article, and (5) collating, summarizing, reporting, and interpreting the results. We reported our process according to the PRISMA Extension for Scoping Reviews (Tricco et al. 2018).

### Research questions

1. How are competency frameworks developed in healthcare professions?
2. How do competency framework development processes align with previous guidance?
3. What insights can be gleaned from the activities of health professions in their developmental activities and their alignment or not with previous guidance?

### Identify relevant studies

#### Systematic search

We structured searches using terms that addressed the development of competency frameworks in healthcare professions. In addition, we considered other related concepts, and combinations of keywords and subject headings that were used are outlined in Appendix I. We selected six databases to ensure a broad range of disciplines were included: MEDLINE, CINAHL, PsycINFO, EMBASE, Scopus, and Education Resources Information Center (ERIC). We also searched grey literature sites greylit.org and Trove, and we reviewed the first 1,000 records from Google Scholar. We title screened citations within articles if they appeared relevant to the review (Greenhalgh and Peacock 2005). Our search was restricted to articles published in English. No limits were set on publication date, study design or country of origin. We conducted pilot searches in May and June 2018 with the help of two information specialists to refine and finalize the search strategy, and we conducted the final searches in August 2018.

Citations were imported into EndNote X8 (Clarivate Analytics, Philadelphia, PA) and we manually removed duplicate citations. The remaining articles were uploaded to the online systematic-review software Covidence (Veritas Health Innovation, Melbourne, Australia) for title and abstract screening, and data characterisation.

### Select the studies

#### Eligibility criteria

Studies were eligible for inclusion if they involved a healthcare profession, produced a competency framework, and explicitly described the development process. Where the same data were reported in more than one publication (e.g., a journal article and a thesis), we only included the version that reported the most complete data. Studies of all types were included.

#### Title and abstract screening

Initial screening comprised of a review of title and abstracts by two reviewers (AB and BW). Disagreements were resolved through discussion until consensus was achieved. Where disagreement remained or there was insufficient evidence to make a decision, the citation was included for full article review.

#### Critical appraisal

In line with the scoping review framework, we did not conduct a critical appraisal (Arksey and O’Malley 2005).

### Chart the data

To support the full-text review, we developed a standardised data extraction form to organize information, confirm relevance, and to extract study characteristics (See Appendix II) (Ritchie and Spencer 2002). The information we collected included study characteristics, objectives of studies, and citations. Relevance was confirmed by sampling population and objectives. Characteristics collected via this form included: Author (year), country; Sampling population; Objective/Aim; Methods used; Count of methods; Outcomes. Additional coding was performed in September 2019 based on peer-review, and included: Rationale; Rationale for methods; Triangulation; Funding. We compiled all data into a single spreadsheet in Microsoft Excel 2013 (Microsoft, Redmond, WA) for coding and analysis.

#### Data summary

Due to variations in terminology, methods and strategies used it was necessary to merge some of these in order to facilitate synthesis. This was an iterative process whereby we reduced variation to produce a discrete list of codes, while retaining the pertinent information in each study. For example, we considered ‘steering groups’, ‘working groups’, ‘committees’, and ‘expert panels’ sufficiently similar to be coded as a form of ‘group technique’, while we coded Delphi process and nominal group technique (NGT) as forms of ‘consensus methods’. Stakeholder deliberation included conferences or workshops (but these may also have been used for other purposes), and alternative strategies including input from professional associations. Codes and their definitions are outlined in Table 2.

**Table 2.** Codes and definitions

| **Code** | **Definition/description of approach** | **Example from included studies** |
| --- | --- | --- |
| **1. Group techniques** | Various methods of group processes were used to draft initial frameworks, edit drafts created by expert panels, or provide expert review and input into framework development. These included steering/working groups, expert groups, and conferences or workshops. | "An Expert Working Group, comprising an SBN [*specialist breast nurse*] and researchers and academics in the field of breast cancer nursing, was convened to synthesise the data that emerged from stakeholder consultation with the published literature." (Yates et al. 2007) |
| 1.1 Conference or workshop | A method whereby events are hosted to bring together small and large groups, of the same or multiple disciplines, in order to gain input, and depending on the format of the event, consensus and feedback to refine and revise the competency framework | "A one-day conference was held with two leaders from each of 31 medical schools" (Z. Liu et al. 2016) |
| **2. Literature review** | A method that seeks to identify current knowledge of a topic, allow for consolidation, and facilitate researchers to build on previous work, to avoid duplication and to identify any omissions or gaps in the knowledge of a topic (Grant and Booth 2009). A review of related research literature was conducted prior to or during the development of the competency framework. These included systematic, scoping, integrative and focused reviews, and other variations such as environmental scans. | “A literature search was carried out in order to identify existing competency frameworks for dementia care” (Smythe et al. 2014) |
| **3. Stakeholder deliberation** | A strategy whereby stakeholders (practitioners, external organisations, experts in the field or other relevant parties) were solicited for feedback | “The professional association, occupational health nurses who were not members of ACORN, and other stakeholders in the workplace were invited to provide feedback.” (Davey 1995) |
| 3.1 Stakeholder deliberation (patient/carer) | A strategy whereby patients and/or their carers were directly engaged in the framework development process as a stakeholder | “A meeting convened in 2010 involved stakeholders in UK nursing education, practice and management, including patient representatives” (Kirk et al. 2014) |
| **4. Mapping exercise** | The competency framework was mapped against existing national or international standards for the profession, or for the health service of the country or region to ensure alignment with existing standards and policies. | “Using the results of the literature review and environmental scan, the Working Group adapted the Irish Palliative Care Competence Framework” (McCallum et al. 2018) |
| **5. Consensus methods** | Methods that involved the input of individuals in order to gain consensus or agreement on items, framework construct or validate a draft framework. Methods included Delphi process, NGT, and group priority sort. |  |
| 5.1 Delphi process | A facilitated group consensus method conducted over several iterations, in order to elicit opinion and response from a panel of experts, with an emphasis on informed judgement (Brown 1968) in order to inform framework development. | “We convened an inter-professional panel of experts to validate our competency framework by way of a modified Delphi technique” (Moaveni et al. 2010) |
| 5.2 Nominal group technique | A method involving a highly structured meeting during which a group of diverse, but representative participants individually respond to questions, present their responses one at a time, and subsequently prioritize responses in an anonymous fashion (Delbecq and Van de Ven 1971) in order to inform framework development | “We employed three methods in this study: (1) nominal group technique (NGT), (2) expert committee, and (3) text analysis” (Ho et al. 2011) |
| 5.3 Group priority sort | A method whereby a group of diverse stakeholders identify priorities, define a complex concept and contribute to informed decision making in topics where evidence is lacking or inappropriate (Jacobson et al. 2013) | “The group priority sort method, which involves engaging groups of stakeholders in sorting and ranking activities, was used as a validation method” (Ling et al. 2017) |
| **6. Survey** | A method whereby an online or offline questionnaire was sent to stakeholders (practitioners, external organisations, experts in the field or other relevant parties) | “...validate the resulting framework by conducting a complex survey of two cohorts of Fellows” (J. R. Frank 2005) |
| **7. Focus groups** | A method that involves in-depth group interviews on a particular topic in which participants are selected because they are a purposive sampling of a specific population (Lederman 1990, p117). Competency frameworks were discussed in focus groups with stakeholders or practitioners. | “Preliminary input was obtained through a series of focus groups” (Myers et al. 2015) |
| **8. Interviews** | A method whereby competency frameworks were discussed in interviews (structured, semi-structured , in-depth, group* etc.) with stakeholders or practitioners | “A purposive sample of 25 nurse leaders provided information during structured interviews” (Amendola 2008) |
| **9. Practice analysis** | A strategy whereby authors sought to directly or indirectly observe the conduct of practice in the context of the practice environment, operationalized through several different methods | “Over 800 hours of specialist critical care nursing practice were observed and grouped into `domains' or major themes of specialist practice” (Dunn et al. 2000) |

#### Data synthesis

We further explored the outlined codes in order to provide insight into their purpose and how they were operationalized. After synthesizing the results, we then organized them by frequency of use from most to least common. We outlined variations that existed within each code including form, function, application, and intended outcomes. This qualitative approach to analysis was performed inductively and iteratively, allowing the data to be representative of itself. The synthesis and subsequent discussion are influenced by our perspective that context is important both in the original studies, and in our own interpretation of the literature. Additionally, study authors may have held underlying positions that are distinct from ours, which may result in differing interpretations of their studies.

## Results

### Search results and study selection

The search yielded 5,669 citations. We identified an additional 110 citations through searches of grey literature, and hand searching. After elimination of duplicates, we screened 5,710 citations at the title and abstract level. This led to the exclusion of 5,331 citations. After full-text review of 379 citations, we included 190 full-texts for data extraction and analysis. See Figure 1 for an illustration of these findings using PRISMA Diagram, and Appendix III for a full list of included studies.

**Figure 1.**
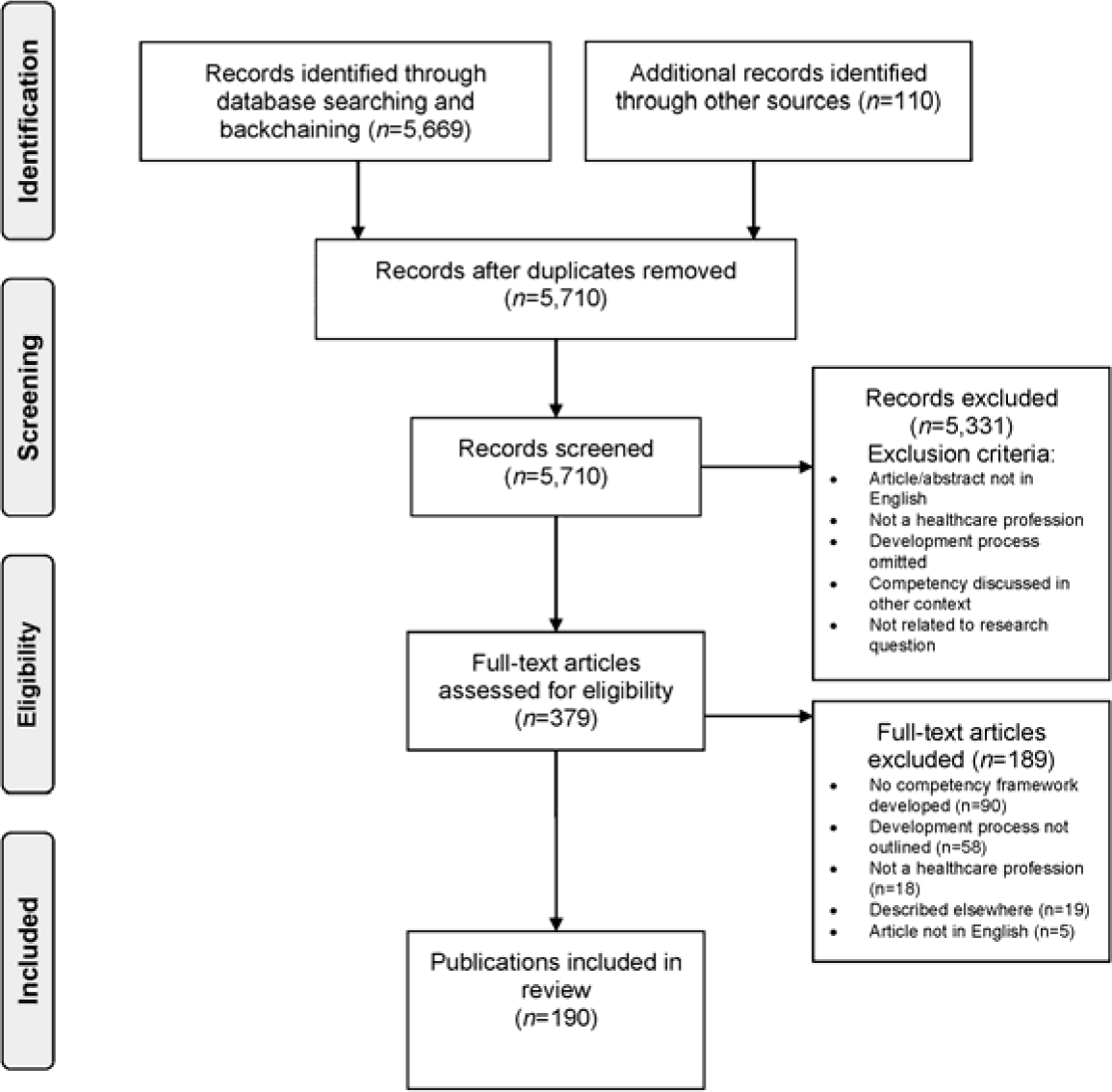
PRISMA Diagram.

### Characteristics of included studies

Included studies were published between 1978 and 2018. The majority were published as peer-reviewed articles (n=172), with the remaining literature comprised of reports (n=13) and theses (n=5). The majority of studies were from the USA (n=65, 34%), followed by the United Kingdom, Canada, and Australia (n=27 each, 14% each). Nursing and medicine competency frameworks accounted for the majority (n=65, 34% each), followed by multidisciplinary frameworks (n=36, 19%). See Table 3 for further characteristics of included studies.

**Table 3.** Study locations and professions.

| **Country** | **n** | **%** |
| --- | --- | --- |
| USA | 65 | 35% |
| UK | 27 | 14% |
| Canada | 27 | 14% |
| Australia | 27 | 14% |
| Brazil | 6 | 3% |
| Europe | 6 | 3% |
| China | 5 | 3% |
| Netherlands | 3 | 2% |
| Global | 3 | 2% |
| South Africa | 2 | 1% |
| Germany | 2 | 1% |
| Lebanon | 2 | 1% |
| India | 2 | 1% |
| Sweden | 1 | 1% |
| Italy | 1 | 1% |
| Thailand | 1 | 1% |
| Korea | 1 | 1% |
| Finland | 1 | 1% |
| Cyprus | 1 | 1% |
| Taiwan | 1 | 1% |
| Mexico | 1 | 1% |
| Western Pacific | 1 | 1% |
| Belgium | 1 | 1% |
| Saudi Arabia | 1 | 1% |
| Africa | 1 | 1% |
| Ireland | 1 | 1% |
| **Profession** | **n** | **%** |
| Nursing | 65 | 34% |
| Medicine | 65 | 34% |
| Multidisciplinary | 36 | 19% |
| Allied health | 10 | 5% |
| Pharmacy | 7 | 4% |
| EMS | 5 | 3% |
| CAM | 1 | 1% |
| Psychology | 1 | 1% |

Literature reviews and group techniques were utilised in 116 studies each (61%). Strategies of stakeholder involvement were utilised in 85 studies (45%), and mapping exercises were conducted in 73 (38%). See Table 4 and Figure 2 for frequency of the methods used, and Appendix IV for full details.

**Table 4.** Frequency of reported methods and strategies

| Method/strategy | n | % |
| --- | --- | --- |
| **1. Group techniques** | 116 | 61% |
| 1.1 Conference / workshop | 40 | 21% |
| **2. Literature review** | 116 | 61% |
| **3. Stakeholder deliberation** | 85 | 45% |
| 3.1 Stakeholder deliberation (patient/carer) | 21 | 11% |
| **4. Mapping exercise** | 73 | 38% |
| **5. Consensus methods** | 54 | 28% |
| 5.1 Delphi process | 49 | 26% |
| 5.2 NGT | 5 | 3% |
| 5.3 Group priority sort | 1 | 0.5% |
| **6. Survey** | 49 | 26% |
| **7. Focus Groups** | 36 | 19% |
| **8. Interviews** | 31 | 16% |
| **9. Practice analysis** | 22 | 12% |

**Figure 2.**
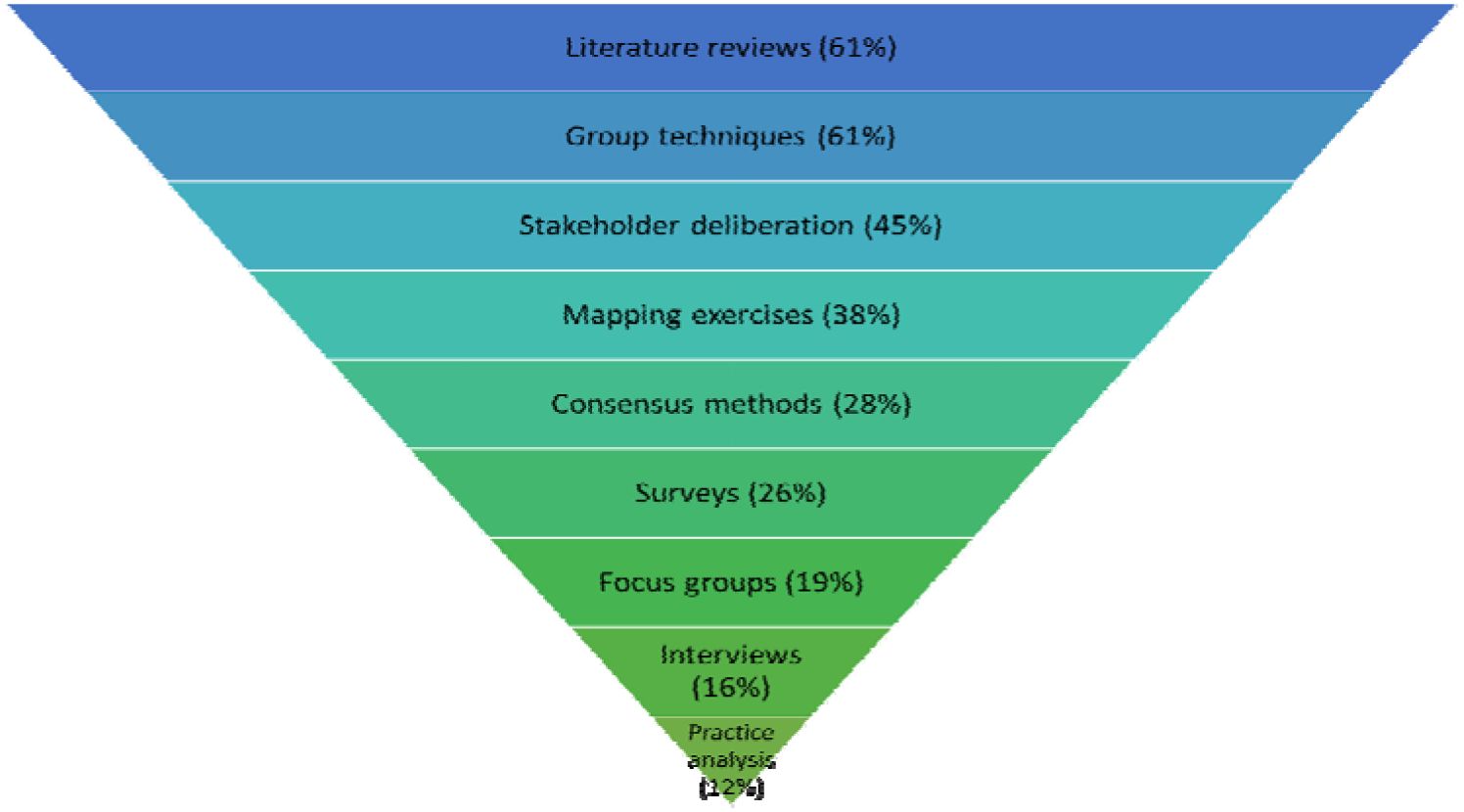
Frequency of reported methods and strategies.

Studies varied in the number of approaches used from one (n=20, 11%) to seven (n=3, 2%) (See Table 5). The median number used was three, and a total of 132 studies (69%) utilised three or more methods or strategies. Combinations of methods varied, and no distinct pattern of use emerged when analysed by profession, location or year. Triangulation of methods was mentioned in 18 (9%) studies. Study periods were not outlined in the majority of studies, but of those that did (n=81, 43%), the timeframe for development ranged from two days to six years, with 41 of these (51%) completed in 12 months or less.

**Table 5.** Number of methods and strategies used by studies

| Number of methods/strategies | n | % |
| --- | --- | --- |
| 1 | 20 | 11% |
| 2 | 39 | 21% |
| 3 | 56 | 29% |
| 4 | 43 | 23% |
| 5 | 26 | 14% |
| 6 | 3 | 2% |
| 7 | 3 | 2% |

All included studies provided a rationale for the development of the framework. Improvement in education was the most commonly reported rationale (medicine), followed by care improvement (nursing) (see Table 6 and Appendix V for more detail). A total of 79 studies (42%) provided a clearly outlined rationale for their choice of methods, and a further 27 (14%) provided a partial rationale. While a detailed analysis of evaluation was outside of the scope of our review, evaluation of the final framework was reported in seven studies (4%), while a further 66 (35%) recommended or planned evaluation. Funding sources were outlined for 110 studies (58%).

**Table 6.**
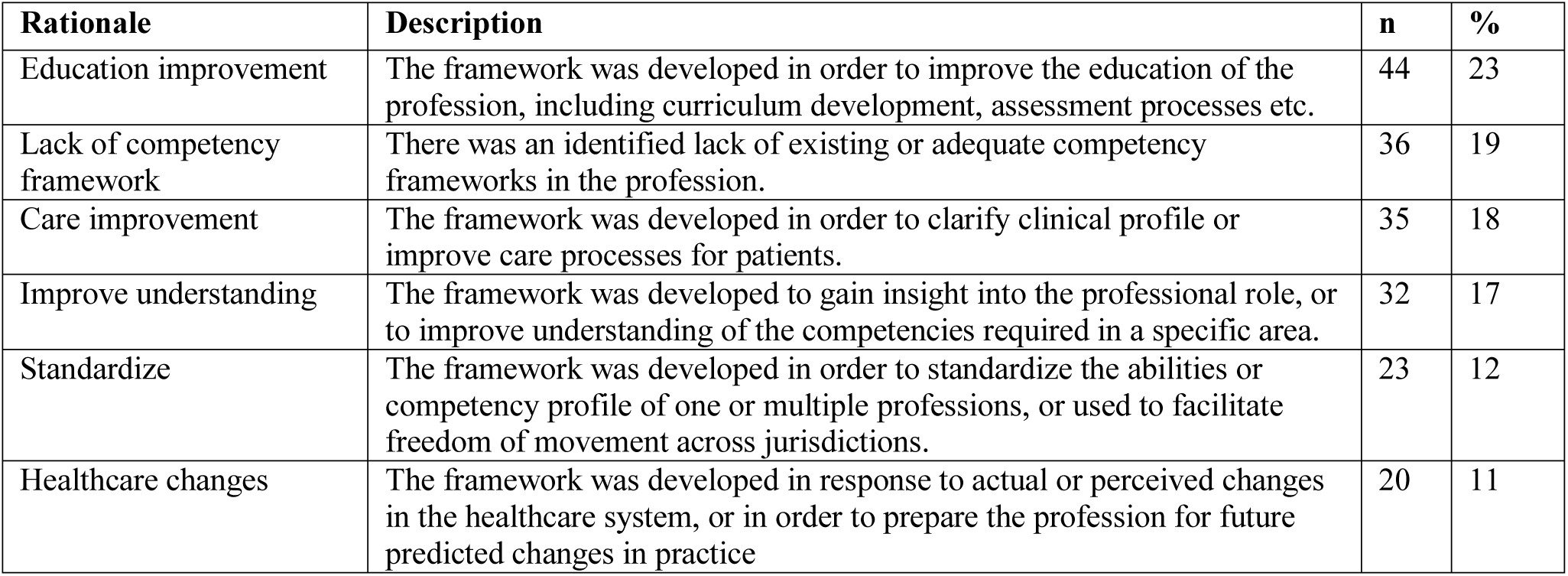
Rationale for development

### Variation in application of approaches

While diversity existed in the methods and strategies used in the development of competency frameworks, we also observed variability within these approaches in form, function, application and intended outcomes (e.g., to achieve consensus, to facilitate dissemination, to review drafts etc.). The variation evident within these approaches suggests that authors made choices (that were not always explicit) in what they valued as meaningful when using such techniques. As such, the functional alignment of these choices remains unclear, and poses a challenge when we attempt to infer alignment with framework objectives. Next we elaborate on these findings, with the exception of DACUM due to its low popularity. Examples are referenced to illustrate variance, but are not intended to be exhaustive lists. See Appendix III for full details.

### Group techniques

Group techniques included working/steering, or expert groups (Aylward et al. 2014; D. Davis et al. 2005), and various group data collection strategies (e.g., conferences and workshops) (Klick et al. 2014; Skirton et al. 2010). Aylward et al. (2014) used a group technique to draft the initial framework, while Davis et al. (2005) used it to edit a draft created by an expert group. Authors including Klick et al. (2014) used a large conference to facilitate input and dissemination. Conversely, others such as Skirton et al. (2010) elected for a smaller group workshop to review drafts and gain input. While there is variation within this category, the same holds true for other sources of evidence for competency framework development.

### Literature reviews

Different types of literature reviews included (a) systematic reviews (Galbraith et al. 2017; Klick et al. 2014), (b) scoping reviews (AlShammari et al. 2018; Redwood-Campbell et al. 2011), (c) integrative reviews (Camelo 2012), (d) focused reviews (Tavares et al. 2016; Yates et al. 2007), and (e) environmental scans (McCallum et al. 2018; National Physiotherapy Advisory Group 2017). Many authors did not explicitly outline the type of review they conducted, and instead described it using generic terms such as ‘broad’, ‘extensive’, and ‘comprehensive’. Some authors performed a review to identify existing competencies (Chen et al. 2013; Hemalatha and Shakuntala 2018), while others performed it to inform subsequent methodology (Davey 1995; Sherbino et al. 2014). It appears that authors make decisions regarding the type, role and relevance of reviews, and integrate them (or not) for a variety of reasons which are often unclear and remain implicit.

### Stakeholder deliberation

We also noted variations in the role and relevance of stakeholder deliberation strategies, which included involving (a) healthcare professionals (J. R. Frank 2005; Yates et al. 2007), (b) professional associations (Davey 1995; Gillan et al. 2013), (c) academics (du Toit et al. 2010; Tangayi et al. 2011), (d) charities and non-profit organisations (Tsaroucha et al. 2013), (e) regulatory bodies (du Toit et al. 2010), (f) trade unions and employers (Reetoo et al. 2005), and (g) patients and their families (R. Davis et al. 2008; Dewing and Traynor 2005). Authors elected to use focus groups (Hamburger et al. 2015), interviews (Tsaroucha et al. 2013), surveys (Tangayi et al. 2011), action research (Dewing and Traynor 2005), conferences and workshops (D. Davis et al. 2005), online wikis (Ambuel et al. 2011), and/or patient advocacy organisations (Yates et al. 2007). Stakeholder input served different purposes, and was used to draft the initial framework (Kirk et al. 2014; Macmillan Cancer Support 2017), to refine and revise draft frameworks (Aylward et al. 2014; Davey 1995), and to gain consensus for the adoption of frameworks (Brewer and Jones 2013; Santy et al. 2005). Despite the focus on ‘patient-centred’ care described in many frameworks, only 21 studies (11%) reported engaging patients or their caregivers. In other instances, it was difficult to understand the role of stakeholders. Who to engage as stakeholders, how to engage them, and for what purpose is, similar to other approaches, also idiosyncratic and thus difficult to infer alignment with framework goals.

### Mapping exercises

The documents used for mapping exercises included (a) specialty board certification exams or reporting milestones (Cicutto et al. 2017; Klein et al. 2014), (b) national policies and health service agendas (Glanville Geake and Ryder 2009; Mills and Pritchard 2004), (c) relevant frameworks from other countries (L. Liu et al. 2014; McCallum et al. 2018), and (d) international or regional frameworks (Barry 2011; Wölfel et al. 2016). These mapping exercises were used as a foundation for framework development (Boyce et al. 2011; McCallum et al. 2018), to identify a pool of items to use in consensus methods (M. Liu et al. 2007), to generate behavioural items for identified competencies (Aylward et al. 2014) and to organise and tabulate responses from stakeholders (Loke and Fung 2014). There appears to be inconsistent adherence with previous development guidance within this approach in relation to the importance of regional context.

### Consensus methods

Consensus methods included (a) Delphi method (Cappiello et al. 2016; Sousa and Alves 2015), (b) group priority sort (Ling et al. 2017) and, (c) nominal group technique (Kirk et al. 2014; Landzaat et al. 2017). Cappiello et al. (2016) used a Delphi method to gain agreement on competencies early in the development of the framework, while Sousa and Alves (2015) used it as a final step to gain consensus. Kirk et al. (2014) utilised NGT as traditionally described (Delbecq and Van de Ven 1971), while Landzaat et al. (2017) utilised a hybrid of modified Delphi and NGT components. Ling et al. (2017) was the only study to utilise group priority sort method. While consensus is a worthwhile strategy that aligns with previous guidance, the rationale for a given approach over another, the sequence, or application was often unclear, and this poses a challenge when we attempt to examine alignment.

### Surveys

Surveys also varied by method, purpose, and characteristics of survey population. From a methodological perspective, some were conducted online or via e-mail (Barnes et al. 2010; Klick et al. 2014), via post (Bluestein 1993; R. Davis et al. 2008), or using a combination of approaches (Baldwin et al. 2007). In terms of function, surveys were utilised to identify initial competencies (Parkinson’s UK 2016), to elicit feedback during the development process (Smythe et al. 2014), and in the subsequent validation of the framework (Sherbino et al. 2014). Sample sizes varied from 33 (Ketterer et al. 2017) to 18,000 (National Physiotherapy Advisory Group 2017), while response rates varied from 3% (NPAG 2017) to 89% (Z. Liu et al. 2016). Actual number of responses ranged from 20 (Ketterer et al. 2017) to 6,247 (Z. Liu et al. 2016). As evidenced within other methods employed, here too we observed variation in the application and function of surveys.

### Focus groups

Focus groups varied in composition, the size and number of groups, and purpose. For example, the composition for some comprised of members of the same discipline (Halcomb et al. 2017; Palermo et al. 2016), while others saw value in using members from different disciplines (Booth and Courtnell 2012; Gillan et al. 2013) - in direct contrast to the *sine qua non* of focus groups (Lederman 1990). In terms of how this method was used, some used it in the initial identification and drafting of competencies (Booth and Courtnell 2012; Patterson et al. 2000), while others used it to engage stakeholders during the development process (Banfield and Lackie 2009; Smythe et al. 2014). Authors including Myers et al. (2015) used focus groups to validate draft frameworks. The reasons for choices made by developers, and the methodological variation evident in this approach remain unclear and inconsistently reported.

### Interviews

The forms of interviews included (a) semi-structured (Akbar et al. 2005; Daouk-Öyry et al. 2017), (b) structured (Amendola 2008), (c) in-depth (Blanchette 2015; Tavares et al. 2016), (d) group interviews (not focus groups) (Loke and Fung 2014), (e) critical incident (Lewis et al. 2010; McCarthy and Fitzpatrick 2009), and (f) behavioural event interviews (Calhoun et al. 2008; Chen et al. 2013). Participants in interviews included patients and family members (Dijkman et al. 2017; Patterson et al. 2000), academics (Chen et al. 2013; Gardner et al. 2006), and healthcare professionals (Calhoun et al. 2008; Chen et al. 2013). Interviews were conducted to gain expert input (Smythe et al. 2014; Tavares et al. 2016), to gain insight into practice (Dunn et al. 2000; McCarthy and Fitzpatrick 2009), to confirm findings from other methods (i.e. triangulation) (Dunn et al. 2000; Palermo et al. 2017), and to solicit contributions from diverse stakeholders (Kwan et al. 2016). The number of interviews conducted was often not reported, however, several authors provided details on population, technique, and analysis for interviews in their studies (Palermo et al. 2017; Tavares et al. 2016). As with other approaches, who to interview, how, and for what purpose was often not adequately reported, and this presents a challenge when we attempt to evaluate the outcomes.

### Practice analysis

Practice analysis involved methods such as (a) functional analysis (Bench et al. 2003; Palermo et al. 2016), (b) analysis of administrative data (Dressler et al. 2006; Stucky et al. 2010), (c) direct observation of practice (Dewing and Traynor 2005; Underwood et al. 1996), (d) critical incident technique (CIT) (Dunn et al. 2000; Lewis et al. 2010), (e) review of position descriptions (Akbar et al. 2005; Fidler 1997), and (f) task or role analysis (Cattini 1999; Chang et al. 2013). Dressler et al. (2006) identified commonly encountered conditions in billing data, while Stucky et al. (2010) and Shaughnessy et al. (2013) identified commonly recorded diagnostic codes to inform the development of competency frameworks. Dunn et al. (2000), Underwood et al. (1996), and Dewing and Traynor (2005) observed practice in-person, while Patterson et al. (2000) observed video recorded interactions to develop an understanding of practice. Practice analyses were used to inform the initial list of competencies (Dressler et al. 2006; Fidler 1997), as a means of capturing the complexity of practice in context (Dunn et al. 2000; Underwood et al. 1996), to triangulate data from other methods (Lewis et al. 2010; McCarthy and Fitzpatrick 2009), and as a means of validating frameworks (Carrington et al. 2011). Timeframes of data collection also varied significantly, and were not always reported. The variation with this approach was perhaps to be expected given the differences in practice between professions. Despite existing guidance related to the importance of job/practice analysis (Lucia and Lepsinger 1999; Roe 2002), this method was rarely utilised, which obligates us to question why given its ability to explore the complexities of practice.

## Discussion

Competency frameworks serve various roles including outlining characteristics of a competent workforce, facilitating mobility, and analysing or assessing expertise. Given how existing development guidelines may be limited, combined with the known complexities of practice and practical challenges faced by framework developers, we sought to understand the choices made when developing competency frameworks. After we examined frameworks across multiple contexts, we suggest that: variability exists in what methods or combinations of methods developers use as well as within methods; there is inconsistent adherence to existing guidance (e.g., most neglect practice analyses, but include multiple methods); limited connections are made between intended use and methodological choices; and, outcomes are inconsistently reported.

Given how competency frameworks are developed, we identified a lack of guidance on how to identify the most appropriate methods. While existing guidance permits and/or encourages a certain flexibility (Table 1), we did not identify any guidance regarding making those choices or examining their suitability for the intended purpose or claims authors intend to make about their outcomes (i.e., competency framework). In other words, existing guidance acknowledges that what we consider fit for one setting or profession and intended use may not be for another, hence the flexibility and variability (Whiddett and Hollyforde 1999). While this seems necessary, existing guidelines also seem to lack organizing conceptual frameworks. As an example, social sciences and humanities research often include conceptual or theoretical frameworks as means to impose, organize, prioritize or align methodological choices. These validity ideals appear challenged by practicalities when developing frameworks. That is, we assume by the heterogeneity in our findings that methodological choices may have been influenced by practicalities such as available resources, timeframes, and the experience and expertise of developers. Other factors may include the maturity of the profession, the perspectives and mandate of the developer (i.e. who is creating the framework), the consistency of roles within the profession, and the complexity of practice which is enacted within broader social contexts. These influencing factors remained largely implicit. Lacking sufficient guidance on these conceptual and practical issues, the utility and validity associated with the framework becomes less clear, or difficult to examine.

Limitations in guidelines related to methodological choices ultimately leave producers and users struggling to make interpretations regarding suitability, utility and validity of competency frameworks. In developing competency frameworks limited in conceptual, theoretical or “use” alignment, we risk the perpetuation of frameworks that adopt a form of unintended or unwarranted legitimacy. This may subsequently result in the creation of what we could term a ‘false-god’ framework, which refers to an object of afforded high value that is illegitimate or inaccurate in its professed authority or capability (Toussaint 2009). That is, despite these limitations when developing competency frameworks, the outcomes are ‘worshipped’, or treated as legitimate or accurate representations of practice without sufficient conceptual or empirical arguments, derived by the methods used, or in alignment with intended purpose. It has been argued for example that social contexts and the complexities of clinical practice remain largely ignored in current competency frameworks (Bradley et al. 2015). Outcomes (i.e. final products) could perhaps (unknowingly) be prioritised over accurate representations of practice, thus limiting their suitability and utility, and threatening validity arguments. Existing guidance cautions that the more important the intended use of the framework, the more that its validity needs to be assured (Heywood et al. 1992; Knapp and Knapp 1995). If validity is compromised, this ‘false-god’ could exert substantial downstream effects including poor definitions of competence as well as threats to curriculum and assessment frameworks. These implications warrant consideration of improved guidance related to development and evaluation processes.

As a way forward, we may need to revisit and refine guidance surrounding competency framework development to include ways of capturing and/or representing the complexity of practice, borrowing from philosophical guidance included in mixed methods research in order to improve suitability, utility and validity, while also establishing reporting and evaluation principles (see Figure 3 for conceptual framework). First, we may need to include leverage if not obligate affordances of conceptual frameworks that have been associated with systems theory, social contexts, and mixed-methods approaches to research in development guidelines. Doing so may provide developers with kinds of organizing frameworks, including the role of underlying philosophical positions, assumptions, commitments and what counts as evidence of rigour and validity. Second, those developing frameworks should consider three when developing a framework in order to align purpose with process: *“binary/continuum; atomistic/holistic; and, context-specific/context-general”* (Child and Shaw 2019). These arguments require developers to explicitly consider the scope or intended use of the framework (which will inform their validity arguments); the level of granularity (which will inform their methods and alignment); and, the contexts in which the framework may be enacted (which will inform the degree of contextual specificity required in the development process). If we integrate organizing frameworks of these kinds and associated arguments into guidelines, it may lead to better alignment between intended uses, methods and sequences such that they are deemed “fit for purpose”. This shifts the emphasis from what or how many methods were used – since any one method can be aligned with more than one purpose – to the theoretical and functional alignment of methods with the rationale for development and intended uses (Child and Shaw 2019). Until implementation of these types of guidelines, we suggest that interpretation of the utility and validity of outcomes (i.e., competency frameworks) may be more variable or less certain (Arundel et al. 2019; Child and Shaw 2019; Simera et al. 2008).

**Figure 3.**
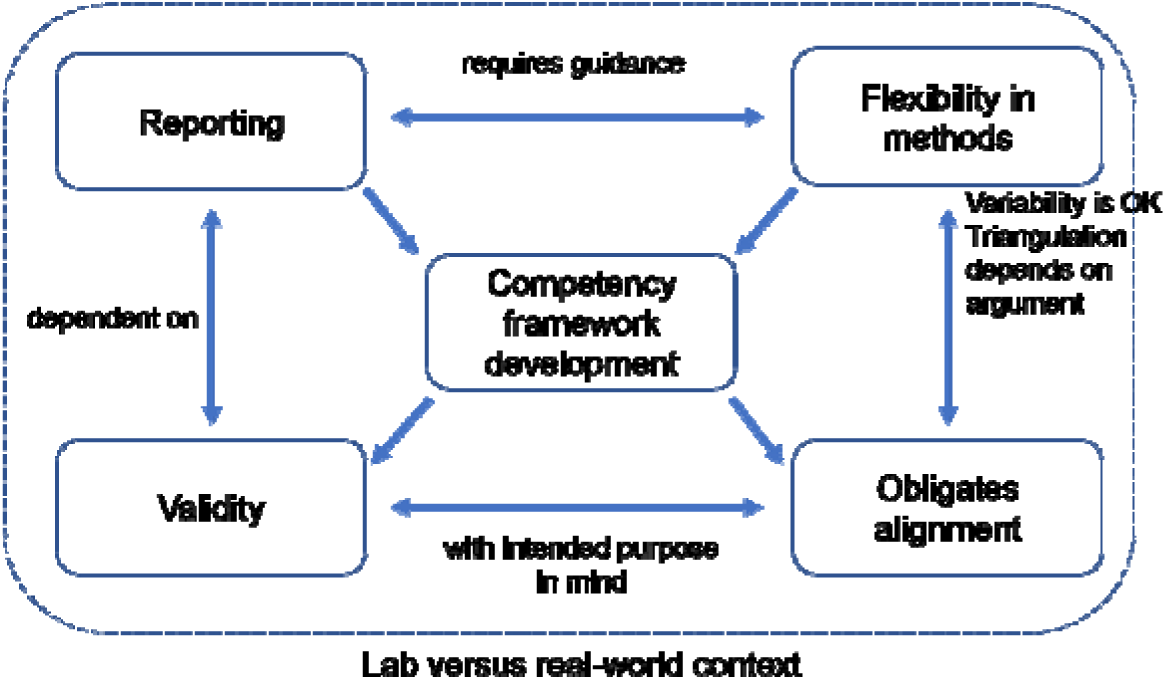
Conceptual framework.

In addition to improved developmental guidelines, we may also support developers and users of competency frameworks through the creation of reporting guidelines that provide structure and clarity (Moher 2007; Moher et al. 2011; Simera et al. 2010). This may include reference to our recommendations above but also incorporate a format that borrows from recently described layered analyses for educational interventions (Cianciolo and Regehr 2019; Horsley and Regehr 2018; Varpio et al. 2012). Applied to competency framework development, techniques may be regarded as surface functions or selected methodologies that are highly context dependent, with underlying principles and philosophy that are context independent. This may help to account for the flexibility required when we attempt to provide guidance to multiple professions across varying contexts. We submit that the suitability, utility and validity of outcomes may leave too much room for interpretation without explicit consideration of the proposals outlined above. However, we acknowledge that the inconsistent adherence to existing guidance we observed in this review suggests that future guidance may also face challenges to implementation.

### Limitations

Our study needs to be considered in the context of its limitations. We may not have identified all relevant studies despite attempts to be comprehensive. While our search strategy included terms previously used to describe the development of competency frameworks in various professions, others may exist. The keywords used to index papers lack consistency and a wide variety of descriptive terms are used in abstracts. Our search and review was restricted to articles published in English, but this does not inherently bias a review (Morrison et al. 2012). The Google Scholar search was limited to the first 1000 results; however, the first 200–300 results from Google Scholar are considered adequate for grey literature searches (Haddaway et al. 2015). No new codes were generated after approximately 50 articles were coded, which suggests that the inclusion of additional literature would likely not have influenced the overall findings of our review. Due to the lack of detail provided by many authors regarding their underlying assumptions, rationale, selection, and conduct of methods, our review cannot provide a concrete overview of all aspects of each included study. Finally, the dynamic nature of research into competency frameworks, EPAs, and the general discourse on competency based education may be considered a limitation. However, our review offers a comprehensive overview of the development of competency frameworks to date along with suggestions for future directions and research.

## Data Availability

Data are available on request.

## Conclusion

Our review identified and explored the research pertaining to competency framework development. Research to date has focused predominantly on the framework outcomes, with considerably less attention devoted to the process of development. Our findings demonstrated that the development process varied substantially, across and within professions, in the choice of methods and in the reporting of the process. There is evidence of inconsistent adherence to existing guidance and a suggestion that existing guidelines may be insufficient. This may result in uncertainty regarding the utility and validity of the outcomes, which may lead to unintended or unwarranted legitimacy. In light of our findings, the development process for competency framework development may benefit from improved guidance. This guidance should obligate a focus on organizing conceptual frameworks that promote the functional alignment of methods and strategies with intended uses and contexts. In addition, such guidance should assist developers to determine approaches that may be better positioned to overcome many of the challenges associated with competency framework development, including sufficiently capturing the complexities of practice. Extending existing guidelines in these ways may be complemented with further research on the implementation, reporting, and evaluation of competency frameworks outcomes.

## Acknowledgments

The authors wish to thank Ms. Paula Todd and Ms. Megan Anderson for their valuable insights into the search strategy.

## Notes

### Competing Interest Statement

The authors have declared no competing interest.

### Funding Statement

No external funding was received.

### Author Declarations

All relevant ethical guidelines have been followed and any necessary IRB and/or ethics committee approvals have been obtained.

Any clinical trials involved have been registered with an ICMJE-approved registry such as ClinicalTrials.gov and the trial ID is included in the manuscript.

